# Dentin deproteinization effect on the push out shear bond strength of self-adhesive resin cements

**DOI:** 10.1101/2025.08.25.25334342

**Authors:** Elnaz Shafigh, mohammad amin yektari, siavash abdollahpoor

**Author notes:** (Corresponding author) shafigh elnaz.

## Abstract

**Objective:** Self-adhesive resin cements are easy to use and do not need further preparation but their bonding ability to dentin is somehow questionable.Ddentin preparation with acidic solutions and therefore removing smear layer can increase bond strength of self-adhesive resin cements to dentin. As a matter of fact it is assumed that deproteinization of dentin can increase contact between cement and dentin. In the present study the effect of deproteinization of dentin with different concentrations and times of sodium hypochlorite on the push out bond strength of resin cement to dentin is assessed.

**Materials and methods:** 60 third molar teeth were selected and divided to 4 groups consisting of 5% and 20% concentrations and 2 and 10 minutes of using. Samples were randomly divided into six groups of ten. All samples used size 2 Whitepost FGM fiber posts (FGM, Joinville, SC, Brazil), with a diameter of 1.2 mm. Samples were cemented using dual-cure self-adhesive TheraCem (Bisco, USA), cured for 40 seconds with an LED Plus device from Woodpecker made in China

All the specimens were then mounted in plastic cylinders. After 24 hours they were cut with 0.2 mm thickness diamond disc to two coronal and apical parts. Push out strength test was done with Instron 1122R (Universal Testing Machine).

**Results:** Increasing the concentration and using time of sodium hypochlorite increased push out bond strength.

**Discussion:** Deproteinization not only can increase push out bond strength but also increase time and concentration can elevate bond strength measures.

## Objective

Teeth that undergo root canal treatment lose part of their structure due to decay, which can weaken their physical structure. Therefore, it is important to ensure that these teeth have a good prognosis during restoration.^1^ These teeth can fully function and satisfactorily serve as fixed or removable prosthesis bases. However, special techniques are needed to rebuild these teeth. Typically, due to decay, previous restorations, and root canal treatments, a significant amount of tooth structure is lost. If the crown structure is largely intact and forces are suitable, a simple restoration can be placed within the access cavity.^2^

However, if a significant amount of tooth structure is lost, other measures need to be taken. In the past, metal posts were often used, but due to aesthetic considerations, the use of colored posts such as glass fiber and zirconia, which provide enough retention for the core, has increased. The restoration of root canal-treated teeth with fiber posts has gained popularity in recent years.

Recent systematic studies have shown that fiber posts have a longer overall lifespan (3 to 7 years) compared to metal posts. These studies examined features such as root fractures and post separations from the canal. Additionally, as more attention is paid to aesthetics in dentistry, the use of fiber posts has become routine. Fiber posts not only have aesthetic benefits but are also biocompatible and have suitable physical properties for bonding to tooth structure and core build-up material.^3^

The generation of fiber posts began with the introduction of carbon fiber posts in the early 1990s. These posts offered high strength, compatibility with adhesive techniques, and elasticity similar to dentin. This elasticity similarity reduces stress concentration and prevents root fractures. The downside of carbon fiber posts was their poor aesthetics due to their dark color. With the development of glass and quartz fiber posts, the use of carbon fiber posts has become rare. The advantage of the new generation of fiber posts is their improved aesthetics due to their colorless and translucency, while retaining the advantages of the previous generation.^4^

Today, the use of fiber posts is prioritized due to their similar elasticity to dentin, ability to bond to the tooth, uniform force distribution, and aesthetics in all-ceramic crowns. The most common cause of clinical failure in using fiber posts for endodontically treated teeth is their separation from the dentin. Currently, various cements are used to bond fiber posts to the dentin canal. Fiber posts, along with adhesive cements, can form an integrated structural and mechanical unit with dentin. The weakest link in the post-cement-dentin assembly is the adhesion of the cement to the dentin surface.^5^

The adhesive strength of the cement directly affects the bonding quality, and the quality of the bond formed between the post-cement-dentin is the most important factor in achieving post retention. To avoid debonding and the undesirable results like post and core loss, time and cost wastage for creating new posts and cores, root fractures, and even the need to extract teeth, it is essential to examine the bonding strength of cements in attaching fiber posts to root dentin.^6^

To reduce technique sensitivity and avoid debonding, changes have been made in the formulation of resin cements and their bonding systems. Typically, during the bonding process, an acidic conditioner is used, washed with water, and dried with air. At this stage, there is a possibility of over-drying the demineralized dentin and collapsing the collagen network, which can result in resin not fully penetrating the entire thickness of the demineralized dentin area. Inadequate washing and remaining acid can result in over-etching the dentin. The self-etch bonding system does not require drying the bonding surface, and the etching and priming steps are combined, reducing technique sensitivity.^7^

In self-adhesive resin cements, separate etching, priming, and bonding steps are not required. The adhesion and bonding occur due to the properties of the resin molecules integrated into the cement itself. Despite the ease of use, there have been concerns about the ability of these cements to form a strong bond with dentin. Laboratory studies examining electron microscopy showed that using these cements does not result in complete demineralization, collagen penetration, and smear layer removal, which can reduce bonding strength with dentin. ^8^

Some studies comparing self-adhesive and self-etch resin cements have reported weaker bonding with dentin for the former group. On the other hand, some studies have found no significant difference in the bond strength between fiber posts cemented with different resin cements and dentin. Given the conflicting findings, choosing a resin cement with ideal bond strength to dentin to prevent debonding of fiber posts remains challenging.^9^

The mineral components on the smear layer are effective buffers that raise the pH at the internal surface, preventing dentin demineralization in that area. Recently, researchers have found that preparing dentin before treatment with acidic solutions and subsequently removing the smear layer improves the reaction of self-adhesive resin cement with dentin. However, the results of these studies often depend on the materials used, and the use of phosphoric acid before treatment reduces bond strength due to the presence of collagen fibrils that prevent cement penetration.^10^

Therefore, it seems that deproteinizing dentin by removing collagen increases the contact between self-adhesive cement and dentin. Dentin matrix mainly consists of type I collagen and non-collagenous proteins. Since these fibers have a high affinity for water, adhesive components are prone to degradation over time. As a result, phenomena like water absorption, washing of the resin, and polymer weakening occur. Deproteinizing methods partially degrade the collagen network, changing the dentin surface and modifying its hydrophilic properties, making it more compatible with hydrophobic resins and increasing bonding strength. Some studies have also mentioned that using hypochlorite reduces bond strength. Given the contradictions present, this study aimed to investigate the effect of the concentration and duration of deproteinization with sodium hypochlorite on the push-out bond strength of self-adhesive resin cements to dentin.^11^

## Methods

We commenced the study following our university ethics committee’s approval (Code: IR.AJAUMS.REC.1399.94). Sixty intact human periodontally involved teeth mature maxillary premolars were used. our samples of teeth have collected from surgical department of our faculty and all of patient gave oral consent.the number of samples was calculated by power analysis method. After cleaning the teeth and disinfecting with 0.5% chloramine solution, we stored them at 4°C in distilled water for use within 1months. Similar teeth were selected, Teeth with similar dimensions and no root decay, with a minimum root length of 14 mm varying by a maximum of 0.5 mm in each target, and lacking cracks/fractures or caries.

Teeth are classified in the 6 groups:

### Group Division

Samples were randomly divided into six groups of ten. All samples used size 2 Whitepost FGM fiber posts (FGM, Joinville, SC, Brazil), with a diameter of 1.2 mm. Samples were cemented using dual-cure self-adhesive TheraCem (Bisco, USA), cured for 40 seconds with an LED Plus device from Woodpecker made in China

Group 1 (unrestored): no preparation in the canal cavity after root canal treatment and fiber posts were cemented with resin cement in the canal 10 min later and served as the negative control.

groups 2 - 5 were divided based on sodium hypochlorite interventions:

Group 2: 5% sodium hypochlorite for 2 minutes

Group 3: 5% sodium hypochlorite for 10 minutes

Group 4: 20% sodium hypochlorite for 2 minutes

Group 5: 20% sodium hypochlorite for 10 minutes

Group 6 (immediately restored): Immediate restoration was performed after finishing the root canal treatment. no intervention such as deproteinization was performed, and fiber posts were cemented with resin cement in the canal. This group was considered the positive control group. The only difference between negative and positive control group is duration of time after post space preparation and post cemented.

In each tooth, the crown was cut from the proximal surface perpendicular to the tooth’s longitudinal axis with a diamond disk, leaving a root length of 14 mm from the CEJ area.

### Criteria for Sample Inclusion and Exclusion

Samples were selected from single-canal premolar teeth that were healthy and had a minimum root length of 14 mm. Teeth with short roots, root decay, or fractures or cracks in the root area were excluded from the study.

### Canal Preparation

To prepare the traditional access cavity, we removed the pulp chamber roof, pulp horns, and lingual shoulder of dentin and standardized it with a #4 steel round bur. The teeth underwent endodontic treatment as follows: We used a ProTaper Sx rotary device (Dentsply Miallefer, Ballaigues, Switzerland) to enlarge the orifice and the canal’s coronal third.

The irrigation used in this study was 0.9% saline. (0.9 G of salt (NaCl) per 100 ml of solution, or 9 G per litre.). In the first step Using a #10 K-file, in the working length 1 mm short of the apical foramen, we prepared the roots with the rotary device up to #F3 in association with irrigation solutions. In this stage The standardized irrigation method was performed for teeth using a 5-mL irrigation syringe and a 31-gauge side-vented needle, the quantity and contact time of each solution were standardized to 5 mL of 5.25% NaOCl (ChloraXiD, PPH Cerkamed, Stalowa-Wola, Poland) for 1 min after each file, then 5 mL of 17% EDTA (Pulpdent, Watertown, MA, USA) for 2 min, and finally 5 mL of 5.25% NaOCl for 1 min to remove the smear layer ^12^Canal preparation was done chemomechanically using the step-back technique with MAF 30 and a size 30 MAC for all samples in the radiographic PA films as a control. Canal filling was done using lateral condensation with gutta-percha and AH 26 sealer. After the sealer setting was completed, the post space in each sample was prepared to a length of 10 mm using a size 2 fiber post drill.^13^

Deproteinization method – after acid etching as previously described, deproteinization with 5% and 20 % NaOCl for 2 or 10 minutes (as shown in the groups above) was done, rinsing with water for 10 s, and drying under a stream of compressed air for 10 s.^14^

### Sample Preparation

Deproteinized samples according to the above methods were washed for 10 seconds and then dried with compressed air. Before fiber post insertion the cement was applied in to canal with lentolo rotary and then fiber post 2 Whitepost FGM fiber posts (FGM, Joinville, SC, Brazil)inserted and cured for 40 seconds (according to manufacture instructions),All samples were mounted in cylindrical plastic molds in transparent acrylic. After 24 hours, cross-sectional samples were obtained from the root area containing the post using a 0.20 mm thick diamond disk sections (Hager and Meisinger GmbH, Neuss, Germany), at a speed of 150 rpm under water resulting in two 4 mm samples. To evaluate the adhesion of the posts, each root was sliced into two -. The sectioning started 1 mm below the CEJ: there was a coronal (B), and 6 mm under the CEJ was an apical (A) section with a thickness of 4 mm each. ^15^

### Push-out Test

The push-out tests were performed with a commercial tensile test machine Instron^®^ 5965 (Instron, High Wycombe, Buckinghamshire, UK) with the following parameters: 2.5 mm/min crosshead speed, 5 kN Instron^®^.The forces as a function of the displacement of fiber posts were recorded determined the peak force. force applied from the apical to the coronal side until the fiber post-dentin bond failed.To calculate the bond strength in MPa, the recorded peak force (N) were divided by the area of the bonded surface, which was calculated using the following formula for each sample, Failure was identified by observing the curve drop drawn by the Instron software. The applied force increased gradually, and the device recorded the maximum force at the moment of failure. ^16^

### Data Analysis

The mean and standard deviation were calculated, and data were analyzed. The normal distribution of the data was determined using the Kolmogorov-Smirnov test in SPSS software. version 23.0; SPSS, Chicago, IL, USA) was used. The Kolmogorov-Smirnov test and Shapiro-Wilk test were used to check the distribution of the data. We used one-way ANOVA for bond strength comparison among the six groups and Dunnett’s multiple comparison t-test for pairwise group comparisons

### Ethical principles

indicates that the study adhered to the ethical principles for medical research involving humans, as outlined in the 1975 version of the Declaration of Helsinki, as further revised and updated in 2024.this study was accepted in ethical committee in number of IR.AJAUMS.REC.1399.94

## Results

A total sample size of n = 60 teeth was included in the study. Table 1 shows the mean push-out bond strength of resin cement to coronal dentin at different times and concentrations. According to this table, a statistically significant difference is seen between the six groups. The highest mean deproteinization with sodium hypochlorite for coronal dentin was in the group with 20% concentration and 10 minutes, while the lowest was in the first control group.(p value less than 0/05)

**Table 1.** the mean push-out bond strength of resin cement to coronal dentin at different times and concentrations.

| Group - Coronal Dentin | Numbers | Mean ± STD |
| --- | --- | --- |
| Control 1(unrestored) | 10 | 6.06 ± 1.2 |
| **Control 2(immediately restored)** | 10 | 9 ± 1.3 |
| **Hypochlorite 5%, 2 min** | 10 | 12.16 ± 1.57 |
| **Hypochlorite 20%, 2 min** | 10 | 14.02 ± 1.36 |
| **Hypochlorite 20%, 10 min** | 10 | 14.2 ± 1.75 |
| **Hypochlorite 5%, 10 min** | 10 | 12.3 ± 1.57 |

Table 2 shows the mean push-out bond strength of resin cement to apical dentin at different times and concentrations. According to this table, a statistically significant difference is seen between the six groups. The highest mean deproteinization with sodium hypochlorite for apical dentin was in the group with 20% concentration and 10 minutes, while the lowest was in the first control group.

**Table 2.**
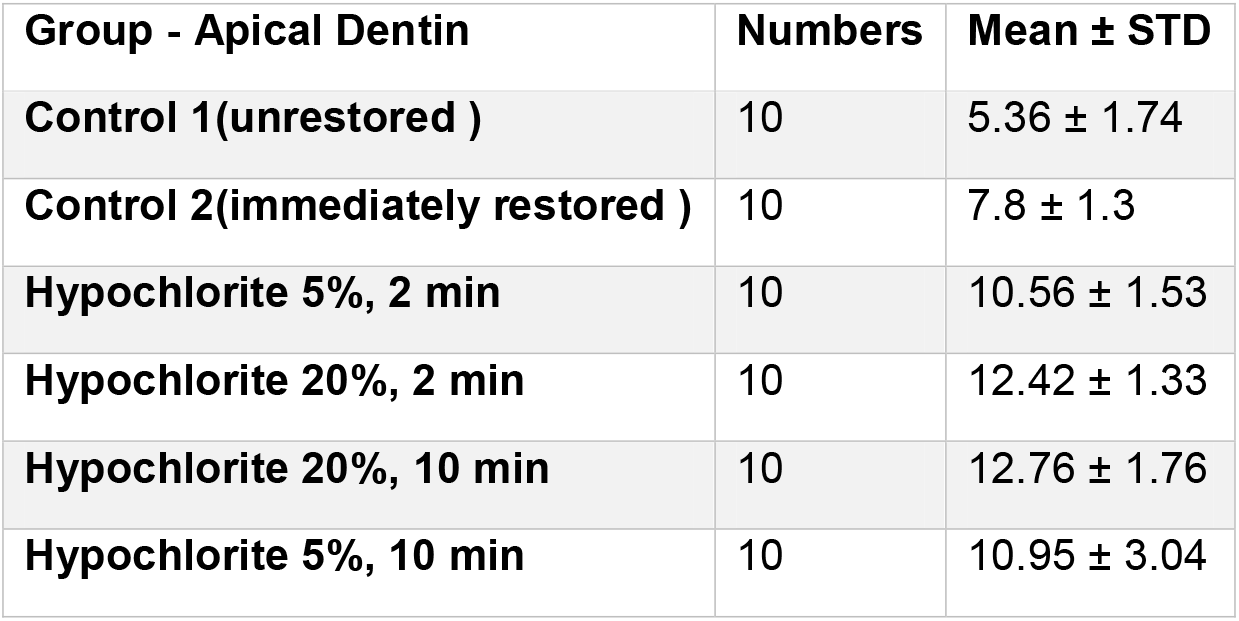
the mean push-out bond strength of resin cement to apical dentin at different times and concentrations.

| **Group - Apical Dentin** | **Numbers** | **Mean ± STD** |
| --- | --- | --- |
| **Control 1(unrestored )** | 10 | 5.36 ± 1.74 |
| **Control 2(immediately restored )** | 10 | 7.8 ± 1.3 |
| **Hypochlorite 5%, 2 min** | 10 | 10.56 ± 1.53 |
| **Hypochlorite 20%, 2 min** | 10 | 12.42 ± 1.33 |
| **Hypochlorite 20%, 10 min** | 10 | 12.76 ± 1.76 |
| **Hypochlorite 5%, 10 min** | 10 | 10.95 ± 3.04 |

Figure 1 shows push-out bond strength of resin cement to apical and coronal dentin at different times and concentrations.

**Figure 1.**
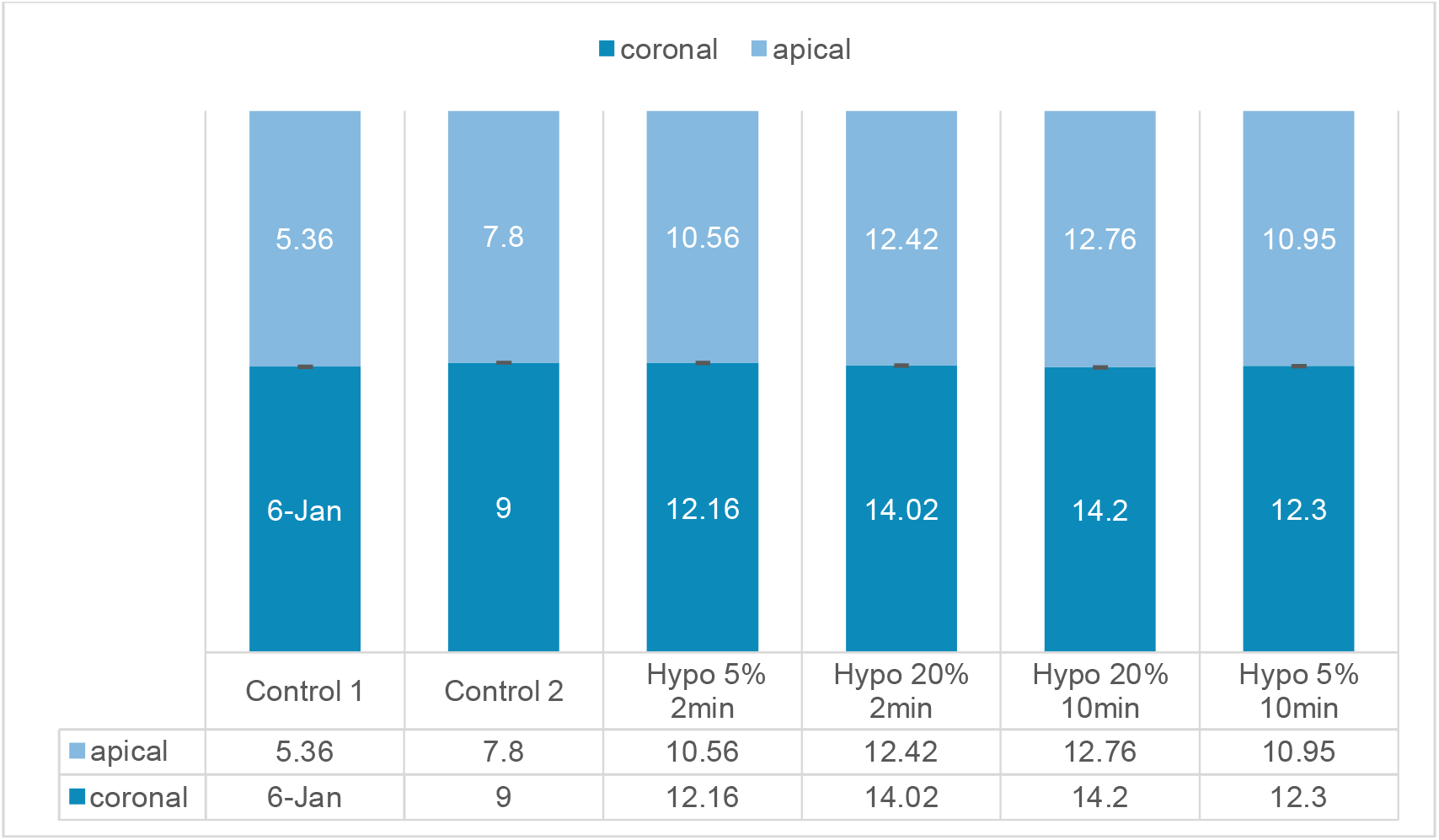
push-out bond strength of resin cement to apical and coronal dentin at different times and concentrations

## Discussion

Today, the use of fiber posts is prioritized due to their similar elasticity to dentin, ability to bond to the tooth, uniform force distribution, and aesthetics in all-ceramic crowns. Past studies on dentin deproteinization before the application of self-etch adhesives and self-adhesive cements have shown contradictory results on shear bond strength. Various studies have been conducted on using fiber posts with different cements and preparations.

^17^Therefore, the current study aimed to help resolve the contradictions in past studies and examine the effect of deproteinization with sodium hypochlorite at different concentrations and durations on the push-out bond strength of self-adhesive resin cements to dentin.

The results of the current study showed that deproteinizing the dentin before applying self-adhesive cement increased the shear bond strength in both apical and coronal regions.

Additionally, increasing the concentration from 5% to 20% also increased the shear bond strength, while increasing the time from 2 to 10 minutes did not have a significant effect on bond strength. According to our study, not only deproteinization increase shear bond strength, but there was also a direct relationship with the concentration of the deproteinizing agent and push out amount.

Examining shear bond strength in the apical and coronal regions, a significant difference was only found in the group with 20% sodium hypochlorite in 10 min, while no significant differences were observed in other groups. This indicates that to achieve higher bond strength in the apical dentin, a duration of 10 minutes should be used, whereas in a single point like apical or coronal dentin alone, no significant difference was observed between 2 minutes and 10 minutes.

In our study results It seems that the concentration of hypochlorite is more important than the duration of deproteinization process, in this manner the use of 20 % hypochlorite for 2 min have greater push out bond strength compared to 5 % in 10 min. recommended that because of time saving hypochlorite 20 % used for 2 min.

In a study by Ehsan Mohammadian Amiri and colleagues in 2017, investigating the effect of self-adhesive and dual-cure cements with separate self-etch adhesives on the bond strength of fiber posts to dentin in different parts of the root, similar studies also used the Instron device to measure mean shear bond strength. In the push-out test, force is applied along the cement-dentin interface, and due to its practical nature, we used this device as well.^18^

As we know, sodium hypochlorite is effective on both the mineral and organic components of dentin, making it highly effective for canal washing in root canal treatment by eliminating bacterial factors. Different studies have investigated the preparation of dentin under the influence of sodium hypochlorite and other substances like chlorhexidine and ethanol or their combinations, with some studies showing a decrease in shear bond strength after using sodium hypochlorite and others showing an increase in bond strength.

Exposing collagen fibers is essential for forming microtags, which play an important role in bond strength due to resin penetration in intertubular dentin. However, over-exposure of collagen fibers can lead to their collapse, preventing the formation of microtags and a strong bond. This could explain why some studies showed a negative impact of sodium hypochlorite on dentin bond strength.

In a study by Shirin Zad, comparing the effect of three canal irrigants on the retention of quartz fiber posts cemented with resin cement, it was concluded that sodium hypochlorite reduced shear bond strength compared to chlorhexidine. Other studies reached similar conclusions, indicating that sodium hypochlorite leads to the oxidation of certain components in the dentin matrix and the formation of radicals derived from protein materials. ^19^These materials compete with the free radicals produced during the light activation of resins, prematurely halting the polymerization process and preventing complete polymerization.

Lisboa also showed that using sodium hypochlorite increased bond strength, which aligns with our study, and attributed this increase to the removal of the smear layer and collagen, resulting in better penetration of the cement between dentin and resin.^20^

Adolson also indicated that using sodium hypochlorite on dentin for bonding fiber posts negatively affects bond strength, showing that this reduction is only observed with self-etch cements, which is consistent with our study. This could be due to different cements being used.^21^

Dentin matrix mainly consists of type I collagen and non-collagenous proteins. Since these fibers have a high affinity for water, adhesive components are prone to degradation over time, leading to water absorption, resin washout, and other phenomena that weaken the polymer^22^. Deproteinizing methods partially degrade the collagen network, changing the dentin surface and modifying its hydrophilic properties, making dentin more compatible with hydrophobic resins. Recent findings indicate that more hydrophobic resin compositions remain stable over time, and deproteinizing also increases the size of resin tags.^23^

## Conclusion

deproteinizing before using self-adhesive cements not only increases shear bond strength but also enhances it with higher concentrations and prolonged durations. Higher concentrations and longer application times enhance the effects of deproteinization, leading to increased shear bond strength when using self-adhesive cements. Because of potential cytotoxic effects of hypochlorite, it recommended more studies done to check out the in vivo toxicity and future clinical applicability.

## Data Availability

All data produced in the present study are available upon reasonable request to the authors

## Author contributions

All authors contributed to the conception, design, data acquisition and interpretation, and statistical analysis and drafted and critically revised the manuscript. All authors gave their final approval and agree to be accountable for all aspects of the work.

## Acknowledgements

The authors express their gratitude to Aja University of Medical Sciences

## Data availability statement

The databases and all of the findings of this study are available on request from the corresponding author. The data are not publicly available because of ethical restrictions.

## References

1 Shafigh, E. and M. Ashrafi, A REVIEW OF MECHANICAL BEHAVIOR OF DENTAL CERAMIC RESTORATIONS. Journal of Mechanics in Medicine and Biology, 2021. 21(08): p. 2150063.

2 Sequeira-Byron P, Fedorowicz Z, Carter B, Nasser M, Alrowaili EF. Single crowns versus conventional fillings for the restoration of root-filled teeth. Cochrane Database Syst Rev. 2015 Sep 25;2015(9):CD009109. doi: 10.1002/14651858.CD009109.pub3. PMID: 26403154; PMCID: PMC7111426.

3 Wang X, Shu X, Zhang Y, Yang B, Jian Y, Zhao K. Evaluation of fiber posts vs metal posts for restoring severely damaged endodontically treated teeth: a systematic review and meta-analysis. Quintessence Int. 2019;50(1):8–20. doi: 10.3290/j.qi.a41499. PMID: 30600326.

4 Joshi S, Shah P, Gandhage D, Mopagar V, Malge RK, Pendyala G. Comparative Evaluation of Fracture Resistance of Carbon Fiber Posts and Glass Fiber Posts in Permanent Anterior Teeth: An In Vitro Study. Cureus. 2024 May 20;16(5):e60647. doi: 10.7759/cureus.60647. PMID: 38903272; PMCID: PMC11187463.

5 Siavash Savadi Oskoee, Mahmoud Bahari, Soodabeh Kimyai, Saeed Asgary, Katayoun Katebi. “Push-out Bond Strength of Fiber Posts to Intraradicular Dentin Using Multimode Adhesive System”, Journal of Endodontics, 2016

6 Ebert J, Leyer A, Günther O, Lohbauer U, Petschelt A, Frankenberger R, Roggendorf MJ. Bond strength of adhesive cements to root canal dentin tested with a novel pull-out approach. J Endod. 2011 Nov;37(11):1558–61. doi: 10.1016/j.joen.2011.08.009. Epub 2011 Sep 15. PMID: 22000463.

7 Perdigão J. Current perspectives on dental adhesion: (1) Dentin adhesion - not there yet. Jpn Dent Sci Rev. 2020 Nov;56(1):190–207. doi: 10.1016/j.jdsr.2020.08.004. Epub 2020 Sep 23. PMID: 34188727; PMCID: PMC8216299.

8 Meharry MR, Schwartz J, Montalvo A, Mueller D, Mitchell JC. Comparison of 2 self-adhesive resin cements with or without a self-etching primer. Gen Dent. 2020 Jan-Feb;68(1):22–28. PMID: 31859658.

9 Amiri EM, Balouch F, Atri F. Effect of Self-Adhesive and Separate Etch Adhesive Dual Cure Resin Cements on the Bond Strength of Fiber Post to Dentin at Different Parts of the Root. J Dent (Tehran). 2017 May;14(3):153–158. PMID: 29167687; PMCID: PMC5694848.

10 Saikaew P, Sattabanasuk V, Harnirattisai C, Chowdhury AFMA, Carvalho R, Sano H. Role of the smear layer in adhesive dentistry and the clinical applications to improve bonding performance. Jpn Dent Sci Rev. 2022 Nov;58:59–66. doi: 10.1016/j.jdsr.2021.12.001. Epub 2022 Jan 29. PMID: 35140823; PMCID: PMC8814382.

11 H S Delgado A, Belmar Da Costa M, Polido MC, Mano Azul A, Sauro S. Collagen-depletion strategies in dentin as alternatives to the hybrid layer concept and their effect on bond strength: a systematic review. Sci Rep. 2022 Jul 29;12(1):13028. doi: 10.1038/s41598-022-17371-0. PMID: 35906302; PMCID: PMC9338246.

12 Özyürek T, Ülker Ö, Demiryürek EÖ, Yılmaz F. The Effects of Endodontic Access Cavity Preparation Design on the Fracture Strength of Endodontically Treated Teeth: Traditional Versus Conservative Preparation. J Endod. 2018 May;44(5):800–805. doi: 10.1016/j.joen.2018.01.020. Epub 2018 Mar 20. PMID: 29571907.

13 Shafiei F, Tavangar MS. Pre-Sealing of Endodontic Access Cavities for the Preservation of Anterior Teeth Fracture Resistance. Clin Exp Dent Res. 2024 Aug;10(4):e936. doi: 10.1002/cre2.936. PMID: 39016080; PMCID: PMC11252827.

14 Lisboa DS, Santos SV, Griza S, Rodrigues JL, Faria-e-Silva AL. Dentin deproteinization effect on bond strength of self-adhesive resin cements. Braz Oral Res. 2013 Jan-Feb;27(1):73–5. doi: 10.1590/s1806-83242013000100013. PMID: 23306629

15 Pereira JR, Rosa RA, Só MV, Afonso D, Kuga MC, Honório HM, Valle AL, Vidotti HA. Push-out bond strength of fiber posts to root dentin using glass ionomer and resin modified glass ionomer cements. J Appl Oral Sci. 2014 Sep-Oct;22(5):390–6. doi: 10.1590/1678-775720130466. Epub 2014 Jul 4. PMID: 25004052; PMCID: PMC4245750.

16 Rajnics, Z.; Pammer, D.; Kőnig-Péter, A.; Turzó, K.; Marada, G.; Radnai, M. Push-Out Bond Strength of Glass Fiber Endodontic Posts with Different Diameters. Materials 2024, 17, 1492. 10.3390/ma17071492

17 Cagidiaco MC, Goracci C, Garcia-Godoy F, Ferrari M. Clinical studies of fiber posts: a literature review. International Journal of Prosthodontics. 2008 Jul 1;21(4).

18 Amiri EM, Balouch F, Atri F. Effect of Self-Adhesive and Separate Etch Adhesive Dual Cure Resin Cements on the Bond Strength of Fiber Post to Dentin at Different Parts of the Root. J Dent (Tehran). 2017 May;14(3):153–158. PMID: 29167687; PMCID: PMC5694848.

19 Morad, Ayda & Taheri, Safoura & Tiznobaik, Azita & Shirinzad, Mehdi & Roshanaei, Ghodratollah & Yavangi, Mahnaz & Fazli, Faezeh. (2024). Investigating the Relationship between Dental Caries and Microbial Plaque with Unknown Infertility: a Case-Control Study. 10.2139/ssrn.4865343.

20 Lisboa DS, Santos SV, Griza S, Rodrigues JL, Faria-e-Silva AL. Dentin deproteinization effect on bond strength of self-adhesive resin cements. Braz Oral Res. 2013 Jan-Feb;27(1):73–5. doi: 10.1590/s1806-83242013000100013. PMID: 23306629.

21 Furuse AY, Cunha LF, Baratto SP, Leonardi DP, Haragushiku GA, Gonzaga CC. Bond strength of fiber-reinforced posts to deproteinized root canal dentin. J Contemp Dent Pract. 2014 Sep 1;15(5):581–6. doi: 10.5005/jp-journals-10024-1583. PMID: 25707830.

22 Shafigh E, Fekrazad R, Bahrani MM. Push-out evaluation on metal and Fiber posts using two different types of cement in a hyper-narrow environment. European Journal of General Dentistry. 2022 Sep;11(03):166–72.

23 Doglas Cecchin, José Flávio Aonso de Almeida, Brenda P.F.A. Gomes, Alexandre Augusto Zaia, Caio Cesar Randi Ferraz. “Inuence of Chlorhexidine and Ethanol on the Bond Strength and Durability of the Adhesion of the Fiber

